# Tandem Lesion Strokes: to stent or not to stent, Insights from a Decade of Experience at a Comprehensive Stroke Center in Argentina

**DOI:** 10.64898/2026.01.12.26343969

**Authors:** Rosales Julieta, González-Aquines Alejandro, Pérez Nicolás, Chasco Manuel, Lopez Matías, Bleise Carlos, Lylyk Iván, Russo María Julieta, Lylyk Pedro

## Abstract

**Background:** Tandem lesion strokes (TLS), defined by the coexistence of an intracranial large- or medium-vessel occlusion and a concomitant cervical internal carotid artery (ICA) stenosis or occlusion, represent a challenging subtype of acute ischemic stroke. Optimal endovascular management remains controversial, particularly regarding the role of emergent carotid artery stenting (eCAS) during mechanical thrombectomy.

**Objective:** To compare the safety and efficacy of emergent carotid artery stenting versus mechanical thrombectomy alone in patients with anterior circulation tandem lesion strokes treated at a comprehensive stroke center.

**Methods:** We conducted a retrospective observational cohort study of consecutive adults with anterior circulation TLS treated with endovascular therapy within 24 hours of symptom onset between January 2015 and July 2025. Patients were categorized into two groups according to treatment strategy: eCAS performed during thrombectomy or mechanical thrombectomy alone (MTa). Primary efficacy outcomes were ordinal shift in 90-day modified Rankin Scale (mRS), excellent outcome (mRS 0–1), and functional independence (mRS 0–2). Secondary efficacy outcome was successful recanalization (TICI ≥2b). Primary safety outcomes included symptomatic intracranial hemorrhage (sICH), in-hospital mortality, and 90-day mortality.

**Results:** A total of 111 patients were included (mean age 71.2 ± 12.6 years; 68.5% male), of whom 74 (67%) underwent eCAS and 37 (33%) received MTa. Patients treated with eCAS achieved higher rates of successful recanalization (97.3% vs 78.4%; OR 13.26, 95% CI 2.13–82.49; *p* = .006) and excellent functional outcomes at 90 days (41.9% vs 12.5%; OR 6.80, 95% CI 1.35–34.20; *p* = .020). There were no significant differences between groups in rates of sICH, early neurological deterioration, or mortality. Ordinal logistic regression showed a non-significant trend toward better functional outcomes with eCAS.

**Conclusions:** In this single-center experience, emergent eCAS during mechanical thrombectomy for TLS was associated with higher reperfusion rates and improved functional outcomes without increased hemorrhagic risk or mortality. These findings support eCAS as a feasible and safe strategy in selected patients and highlight the need for prospective randomized trials.

## BACKGROUND

Tandem lesions strokes (TLS) refer to the simultaneous presence of an intracranial large- or medium-vessel occlusion (LVO/MeVO) and a concomitant stenosis or occlusion of the cervical internal carotid artery (ICA). They account for approximately 10%–20% of all LVO strokes (1). Management of TLS is particularly challenging, as it is associated with lower recanalization rates, poorer functional outcomes, higher rates of severe disability, and increased mortality (2,3). Only 4% to 32% of TLS patients treated with intravenous thrombolysis achieve recanalization, and 17% achieve a good clinical outcome, with a death rate as high as 55% (4).

The lack of a standard approach to treating TLS has resulted in highly variable practices and outcomes (5,6). Some interventionists perform emergent carotid artery stenting (eCAS) during mechanical thrombectomy (MT) to achieve immediate recanalization, either before (anterograde approach) or afterward (retrograde approach) intracranial thrombectomy. Others advocate alternative strategies, such as balloon angioplasty, aspiration of the cervical segment, or performing intracranial MT alone, followed by delayed treatment of the cervical lesion (MTa, mechanical thrombectomy alone). In addition, there is no agreement on the antithrombotic regimen post-CAS, as the potential benefit of reducing ischemic complications from antithrombotic therapy must be balanced against the risk of reperfusion with symptomatic intracranial hemorrhage (sICH) (7).

The challenges in treating TLS highlight a critical gap in knowledge about its optimal endovascular management, largely due to the exclusion of tandem lesions from endovascular thrombectomy trials (8–11). Recent observational studies have suggested potential benefits of eCAS, yet results remain inconsistent across different populations and centers. Although some studies report benefits of eCAS with favorable outcomes (5,6), others observed no differences in successful revascularization, clinical outcomes, or mortality after eCAS (12,13)

Therefore, we aim to assess the safety and efficacy of eCAS compared to the MTa approach in patients presenting with anterior circulation TLS at a single comprehensive stroke center in Argentina, providing contemporary, real-world data from a Latin American cohort.

## METHODS

The methods are reported in accordance with the Strengthening the Reporting of Observational Studies in Epidemiology (STROBE) guidelines (14).

### Study Design, Settings, and Participants

This cross-sectional study used data from a stroke registry from a single comprehensive stroke center in Argentina. The population consisted of consecutive patients with anterior circulation TLS treated with endovascular therapy within 24 hours after symptom onset, between January 1st, 2015, and July 31 th 2025.

Inclusion criteria were an age of 18 years or older, endovascular therapy for intracranial occlusion, and the presence of an extracranial internal carotid artery (ICA) lesion on admission, diagnosed through computed tomography angiography, magnetic resonance angiography, and/or intraprocedural digital subtraction angiography. Patients with isolated extracranial ICA lesions were excluded.

Tandem lesions were defined as an intracranial LVO/MeVO, including petrous, cavernous, or terminus segment of the ICA, M1 to M3 segment of the middle cerebral artery (MCA) and A1 to A3 of the segment of the anterior cerebral artery (ACA) with a concomitant extracranial ICA stenosis of 50% or more and/or occlusion, as defined by the North American Symptomatic Carotid Endarterectomy Trial (NASCET) criteria (15).

This study was approved by the local institutional review board under a waiver of informed consent.

### Study Groups, Data Elements, Exposures, and Interventions

Patients were divided into two groups according to the treatment strategy: (1) eCAS group (patients treated with stenting of the cervical lesion during MT) and (2) MTa group (patients treated with cervical balloon angioplasty or thrombectomy with thromboaspiration and/or stent retriever, only aspiration of the extracranial ICA lesion, or deferred or no extracranial ICA intervention).

From medical records, we collected data on demographic characteristics, vascular risk factors, comorbidities, and NIHSS scores (16) (at admission, at 24 hours and discharge), as well as modified Rankin Scale (mRS) scores (17) (pre-stroke, at discharge, and at 90 days). Antithrombotic therapy was categorized as single/dual antiplatelet therapy, or anticoagulation, and documented for three time points: prior to stroke, during the endovascular procedure, and at discharge. We also recorded the use of intravenous or intra-arterial thrombolysis.

Imaging data included baseline Alberta Stroke Program Early CT Score (ASPECTS) (18) and post-procedural revascularization grade according to the Thrombolysis in Cerebral Infarction score (TICI score) (19). Stroke workflow metrics included onset-to-groin puncture (OTG) times. Anesthesia type was classified as general or neuroleptoanesthesia.

Complications after thrombectomy were recorded, including sICH according to ECAS III criteria (20), ipsilateral ischemic stroke during hospitalization, early neurological deterioration (NIHSS increase ≥4 points within 24 hours after MT), and in-hospital mortality. All intracranial occlusions were treated using a stent retriever and/or contact aspiration catheters, with intracranial stenting or angioplasty performed as needed. The cervical revascularization approach for ICA stenting was categorized as anterograde or retrograde.

### Efficacy and safety Outcomes

The primary efficacy outcomes were ordinal shift in 90-day mRS score, excellent outcome (mRS 0–1) and functional independence (mRS 0–2) at 90 days. The secondary efficacy outcome was successful recanalization, defined as TICI 2b or higher. The primary safety outcomes were sICH, in hospital mortality and mortality at 90 days. Secondary safety outcome was early neurological deterioration.

### Statistical Analysis

Descriptive statistics were used to summarize continuous and categorical variables. We reported categorical variables as numbers (percentages) and continuous variables as means (standard deviations, SDs) or medians (interquartile ranges, IQRs). The Shapiro-Wilk test and histograms were used to assess the normality of distributions. For the bivariate analysis, we used two-tailed paired t-tests or the U Mann-Whitney test for continuous variables based on the distribution, and the χ2 or Fisher exact test for categorical variables, as needed. An ordinal logistic regression model was fitted using mRs at 90 days to identify the direction and magnitude of the association while accounting for the hierarchical nature of mRs. A multivariate logistic regression model was fitted to explore the relationship between a set of independent variables selected for their clinical relevance and prior literature, and sICH and mRs at 90 days. Model calibration was assessed using the Hosmer–Lemeshow goodness-of-fit test, and multicollinearity was evaluated using Variance Inflation Factors (VIFs). A p-value <0.05 was considered statistically significant. The analysis was conducted using SPSS v23

## RESULTS

We included 111 patients with anterior TLS, with a mean age of 71.2 ± 12.55 years and 68.5% men. Of whom, 74 (67%) patients were in the eCAS group and 37 patients (33%) in the MTa group. Compared to the MTa group, patients in the eCAS group presented lower median (IQR) NIHSS at admission (14.5 [IQR 9 - 19] vs 17 [IQR 13.5 - 20]; *p =* .033) and received intra-arterial thrombolysis more frequently (17.6% vs. 2.7%, *p* = .032). In the perioperative setting, the eCAS group received more frequently intravenous tirofiban (52.7% vs 5.4%, *p* < .001) and charge with dual antiplatelet therapy (DAPT) (64.9% vs 16.2%, *p <*.001) and were more often treated with DAPT at discharge (71.2% vs. 29.7%, *p* <.001). The demographic and baseline characteristics of the two groups are presented in **Table 1**.

**Table 1.**
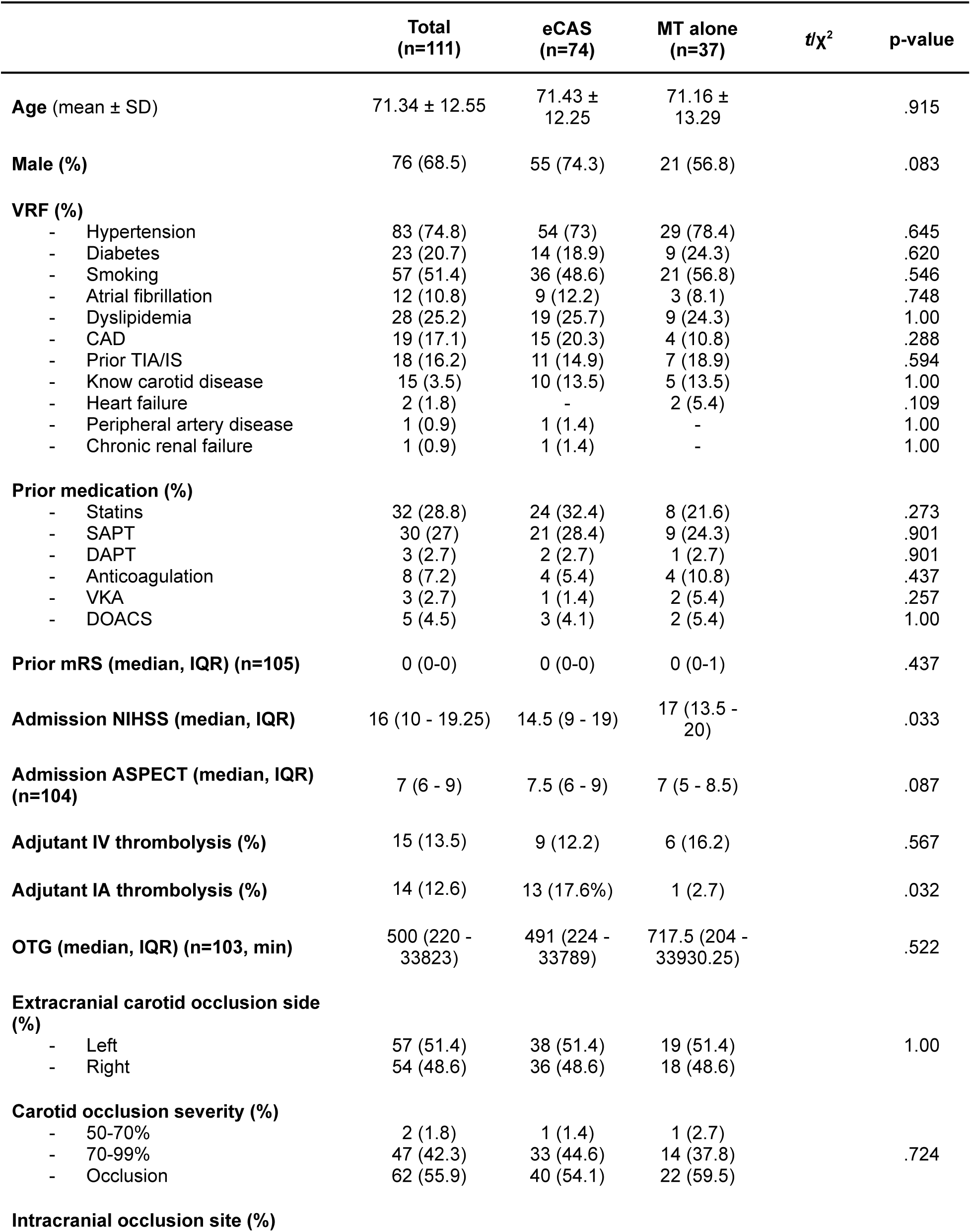
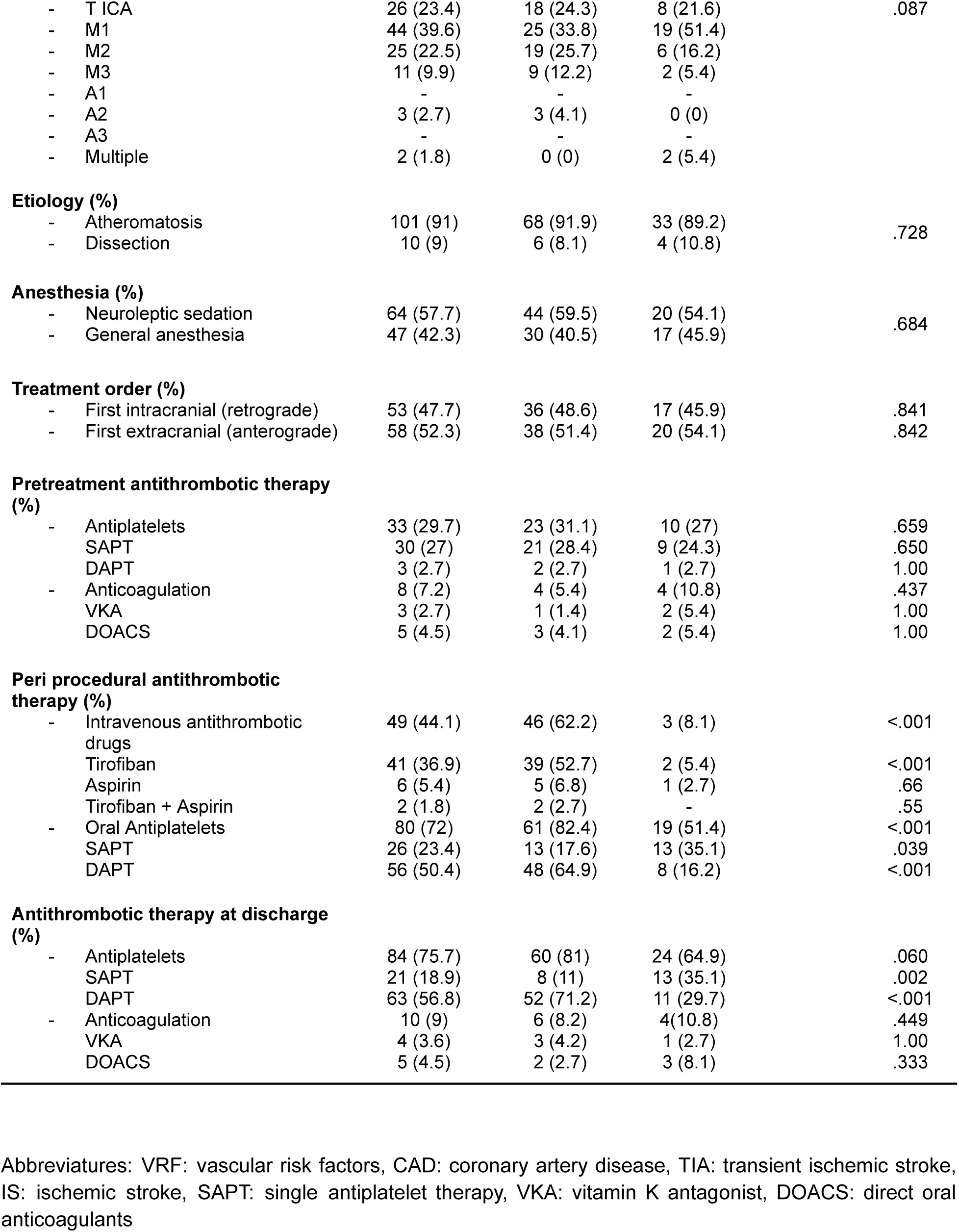
Baseline features of patients included.

|  | **Total**<br>**(n=111)** | **eCAS**<br>**(n=74)** | **MT alone**<br>**(n=37)** | **t/$\chi^2$** | **p-value** |
| --- | --- | --- | --- | --- | --- |
| **Age** (mean ± SD) | 71.34 ± 12.55 | 71.43 ± 12.25 | 71.16 ± 13.29 | | .915 |
| **Male (%)** | 76 (68.5) | 55 (74.3) | 21 (56.8) |  | .083 |
| **VRF (%)** |  |  |  |  |  |
| - Hypertension | 83 (74.8) | 54 (73) | 29 (78.4) |  | .645 |
| - Diabetes | 23 (20.7) | 14 (18.9) | 9 (24.3) |  | .620 |
| - Smoking | 57 (51.4) | 36 (48.6) | 21 (56.8) |  | .546 |
| - Atrial fibrillation | 12 (10.8) | 9 (12.2) | 3 (8.1) |  | .748 |
| - Dyslipidemia | 28 (25.2) | 19 (25.7) | 9 (24.3) |  | 1.00 |
| - CAD | 19 (17.1) | 15 (20.3) | 4 (10.8) |  | .288 |
| - Prior TIA/IS | 18 (16.2) | 11 (14.9) | 7 (18.9) |  | .594 |
| - Know carotid disease | 15 (3.5) | 10 (13.5) | 5 (13.5) |  | 1.00 |
| - Heart failure | 2 (1.8) | - | 2 (5.4) |  | .109 |
| - Peripheral artery disease | 1 (0.9) | 1 (1.4) | - |  | 1.00 |
| - Chronic renal failure | 1 (0.9) | 1 (1.4) | - |  | 1.00 |
| **Prior medication (%)** |  |  |  |  |  |
| - Statins | 32 (28.8) | 24 (32.4) | 8 (21.6) |  | .273 |
| - SAPT | 30 (27) | 21 (28.4) | 9 (24.3) |  | .901 |
| - DAPT | 3 (2.7) | 2 (2.7) | 1 (2.7) |  | .901 |
| - Anticoagulation | 8 (7.2) | 4 (5.4) | 4 (10.8) |  | .437 |
| - VKA | 3 (2.7) | 1 (1.4) | 2 (5.4) |  | .257 |
| - DOACS | 5 (4.5) | 3 (4.1) | 2 (5.4) |  | 1.00 |
| **Prior mRS (median, IQR) (n=105)** | 0 (0-0) | 0 (0-0) | 0 (0-1) |  | .437 |
| **Admission NIHSS (median, IQR)** | 16 (10 - 19.25) | 14.5 (9 - 19) | 17 (13.5 - 20) |  | .033 |
| **Admission ASPECT (median, IQR) (n=104)** | 7 (6 - 9) | 7.5 (6 - 9) | 7 (5 - 8.5) |  | .087 |
| **Adjutant IV thrombolysis (%)** | 15 (13.5) | 9 (12.2) | 6 (16.2) |  | .567 |
| **Adjutant IA thrombolysis (%)** | 14 (12.6) | 13 (17.6%) | 1 (2.7) |  | .032 |
| **OTG (median, IQR) (n=103, min)** | 500 (220 - 33823) | 491 (224 - 33789) | 717.5 (204 - 33930.25) |  | .522 |
| **Extracranial carotid occlusion side (%)** |  |  |  |  |  |
| - Left | 57 (51.4) | 38 (51.4) | 19 (51.4) |  | 1.00 |
| - Right | 54 (48.6) | 36 (48.6) | 18 (48.6) |  |  |
| **Carotid occlusion severity (%)** |  |  |  |  |  |
| - 50-70% | 2 (1.8) | 1 (1.4) | 1 (2.7) |  |  |
| - 70-99% | 47 (42.3) | 33 (44.6) | 14 (37.8) |  | .724 |
| - Occlusion | 62 (55.9) | 40 (54.1) | 22 (59.5) |  |  |
| **Intracranial occlusion site (%)** |  |  |  |  |  |

|  |  |  |  |  |
| --- | --- | --- | --- | --- |
| - T ICA | 26 (23.4) | 18 (24.3) | 8 (21.6) | .087 |
| - M1 | 44 (39.6) | 25 (33.8) | 19 (51.4) |  |
| - M2 | 25 (22.5) | 19 (25.7) | 6 (16.2) |  |
| - M3 | 11 (9.9) | 9 (12.2) | 2 (5.4) |  |
| - A1 | - | - | - |  |
| - A2 | 3 (2.7) | 3 (4.1) | 0 (0) |  |
| - A3 | - | - | - |  |
| - Multiple | 2 (1.8) | 0 (0) | 2 (5.4) |  |
| **Etiology (%)** |  |  |  |  |
| - Atheromatosis | 101 (91) | 68 (91.9) | 33 (89.2) | .728 |
| - Dissection | 10 (9) | 6 (8.1) | 4 (10.8) |  |
| **Anesthesia (%)** |  |  |  |  |
| - Neuroleptic sedation | 64 (57.7) | 44 (59.5) | 20 (54.1) | .684 |
| - General anesthesia | 47 (42.3) | 30 (40.5) | 17 (45.9) |  |
| **Treatment order (%)** |  |  |  |  |
| - First intracranial (retrograde) | 53 (47.7) | 36 (48.6) | 17 (45.9) | .841 |
| - First extracranial (anterograde) | 58 (52.3) | 38 (51.4) | 20 (54.1) | .842 |
| **Pretreatment antithrombotic therapy (%)** |  |  |  |  |
| - Antiplatelets | 33 (29.7) | 23 (31.1) | 10 (27) | .659 |
| SAPT | 30 (27) | 21 (28.4) | 9 (24.3) | .650 |
| DAPT | 3 (2.7) | 2 (2.7) | 1 (2.7) | 1.00 |
| - Anticoagulation | 8 (7.2) | 4 (5.4) | 4 (10.8) | .437 |
| VKA | 3 (2.7) | 1 (1.4) | 2 (5.4) | 1.00 |
| DOACS | 5 (4.5) | 3 (4.1) | 2 (5.4) | 1.00 |
| **Peri procedural antithrombotic therapy (%)** |  |  |  |  |
| - Intravenous antithrombotic drugs | 49 (44.1) | 46 (62.2) | 3 (8.1) | <.001 |
| Tirofiban | 41 (36.9) | 39 (52.7) | 2 (5.4) | <.001 |
| Aspirin | 6 (5.4) | 5 (6.8) | 1 (2.7) | .66 |
| Tirofiban + Aspirin | 2 (1.8) | 2 (2.7) | - | .55 |
| - Oral Antiplatelets | 80 (72) | 61 (82.4) | 19 (51.4) | <.001 |
| SAPT | 26 (23.4) | 13 (17.6) | 13 (35.1) | .039 |
| DAPT | 56 (50.4) | 48 (64.9) | 8 (16.2) | <.001 |
| **Antithrombotic therapy at discharge (%)** |  |  |  |  |
| - Antiplatelets | 84 (75.7) | 60 (81) | 24 (64.9) | .060 |
| SAPT | 21 (18.9) | 8 (11) | 13 (35.1) | .002 |
| DAPT | 63 (56.8) | 52 (71.2) | 11 (29.7) | <.001 |
| - Anticoagulation | 10 (9) | 6 (8.2) | 4 (10.8) | .449 |
| VKA | 4 (3.6) | 3 (4.2) | 1 (2.7) | 1.00 |
| DOACS | 5 (4.5) | 2 (2.7) | 3 (8.1) | .333 |

Abbreviations: VRF: vascular risk factors, CAD: coronary artery disease, TIA: transient ischemic stroke, IS: ischemic stroke, SAPT: single antiplatelet therapy, VKA: vitamin K antagonist, DOACS: direct oral anticoagulants

There was a significantly increased rate of patients with excellent outcome (mRS 0-1) at 90 days in the eCAS group compared with the MTa group (41.9% vs 12.5%; *OR*: 6.80, 95% CI 1.35 - 34.20; *p* = .020); this arm also presented a nonsignificant increase rate in functional independence (mRS 0-2) at 90 days (51.6% vs 32%; *OR*: 1.95, 95% CI 0.62 - 6.17; *p =* .253) and a higher rate of successful recanalization (97.3% vs 78.4%, *OR*: 13.26, 95% CI 2.13 - 82.49; *p =* .006). The rate of sICH was comparable between the two groups (26% vs 24.3%; *OR*: 1.17, 95% CI 0.44 - 3.06; *p* = .755). Similarly, no difference was found in END, in-hospital mortality, and mortality at 90 days. Efficacy and safety outcomes are summarized in **Table 2**.

**Table 2.**
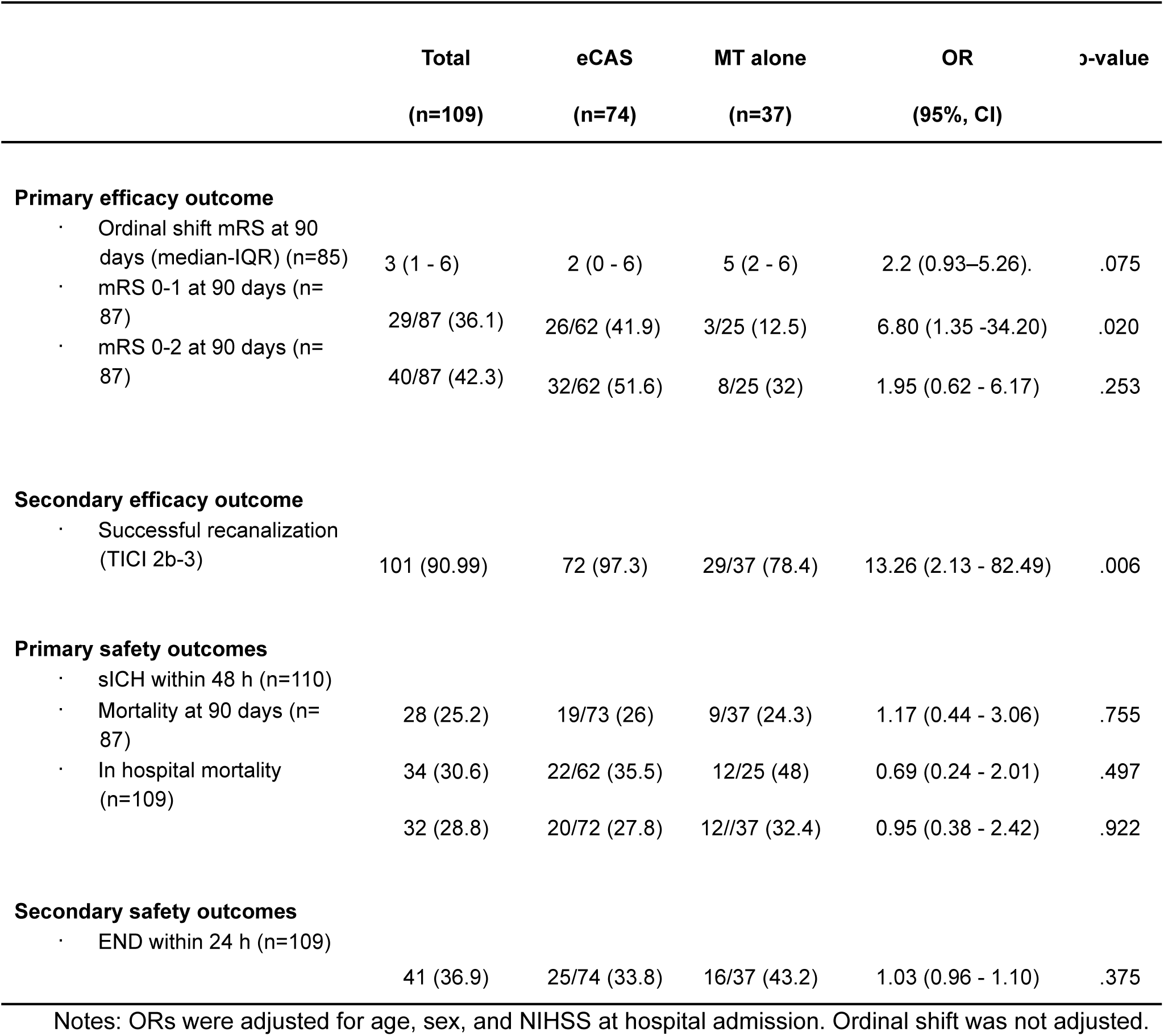
Adjusted bivariate analysis of efficacy and safety outcomes among patients in MTa and eCAS groups.

In an ordinal logistic regression model of 90-day mRS, patients in the eCAS arm tended to have better functional outcomes than those in the MTa arm, although this difference did not reach statistical significance (*OR* for better outcome in eCAS vs MTa ≈ 2.2; 95% CI 0.93–5.26; *p* = .075). Model fit was adequate according to Pearson and Deviance goodness-of-fit tests, and pseudo-R² values indicated that the treatment group explained only a small proportion of the variability in functional outcome.

Finally, a multivariate logistic regression was conducted to identify variables associated with sICH and a good functional outcome at 90 days (mRS 0-2) **(Tables 3 and 4)**. The Hosmer–Lemeshow goodness-of-fit test yielded p > 0.10 for both models., and all VIF values were below 2, indicating no evidence of multicollinearity. Patients not receiving an APT before the event were less likely *(OR* 0.24, CI 95% 0.09 - 0.67; *p =* .006) to develop sICH. Variables associated with a good functional outcome 90 days after the event were age (*OR* 0.93, CI 95% 0.88 - 0.98; *p* = .011) and not having received APT before the event *(OR* 7.12, CI 95% 1.58 - 32.03; p = .01).

**Table 3.**
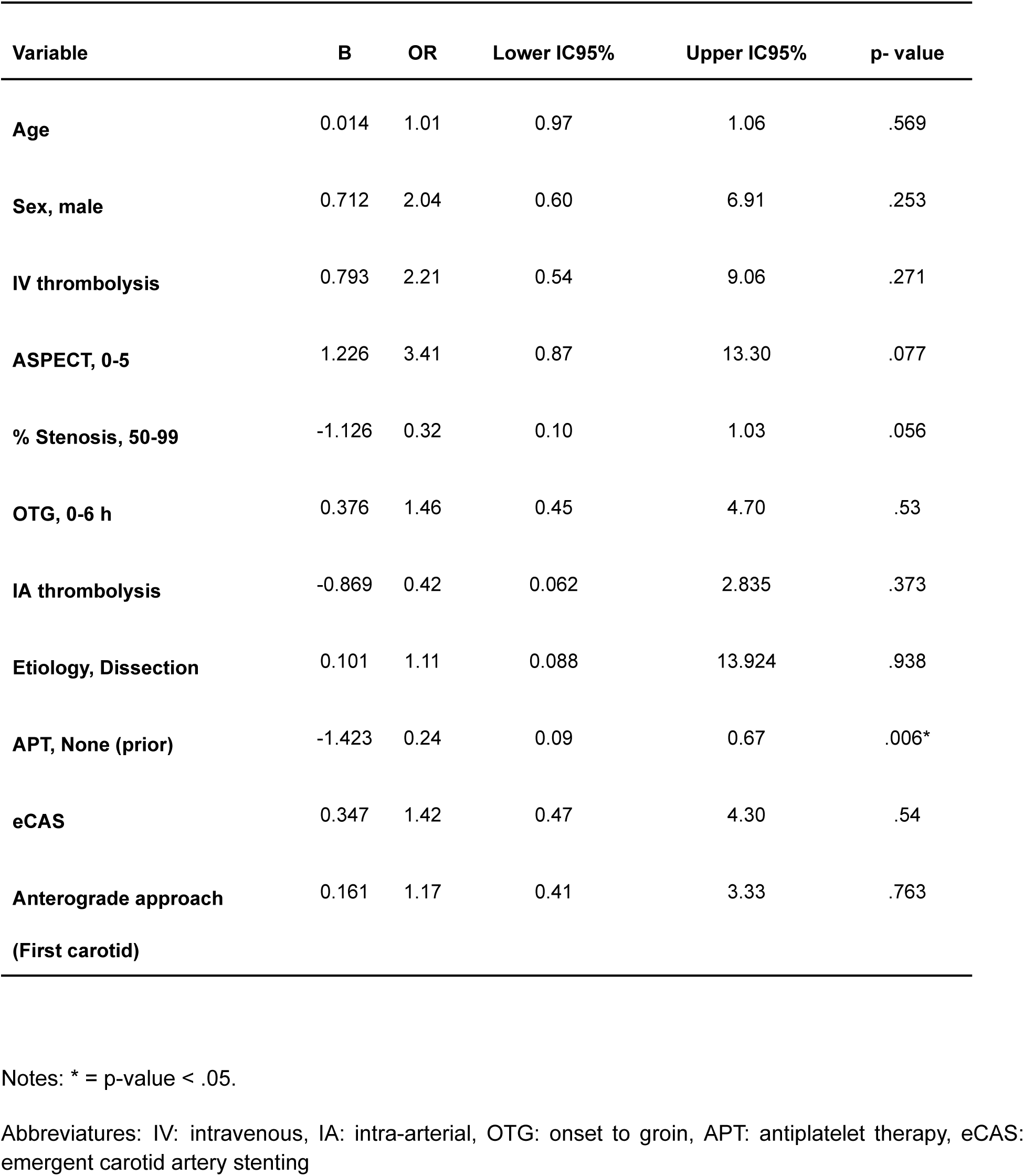
Multivariate logistic regression for sICH.

**Table 4.** Multivariate logistic regression for mRS 0-2 at 90 days.

| Variable | B | OR | Lower IC95% | Upper IC95% | P value |
| --- | --- | --- | --- | --- | --- |
| Age | -0.073 | 0.93 | 0.88 | 0.98 | .011* |
| Sex, male | -0.306 | 0.74 | 0.22 | 2.42 | .614 |
| IV thrombolysis | -0.361 | 0.70 | 0.14 | 3.50 | .661 |
| ASPECT, 0-5 | -1.946 | 0.14 | 0.02 | 1.17 | .069 |
| % Stenosis, 50-99 | 0.469 | 1.60 | 0.51 | 4.99 | .419 |
| OTG, 0-6 hrs | 0.835 | 2.30 | 0.61 | 8.64 | .215 |
| IA thrombolysis | 1.262 | 3.53 | 0.66 | 18.76 | .138 |
| Etiology, Dissection | -0.14 | 0.87 | 0.07 | 10.49 | .912 |
| APT, None (prior) | 1.963 | 7.12 | 1.58 | 32.03 | .01* |
| eCAS | 0.714 | 2.04 | 0.52 | 7.95 | .303 |
| Anterograde approach<br>(First carotid) | -0.984 | 0.37 | 0.11 | 1.23 | .106 |

Notes: \* = p-value < .05.
Abbreviations: OTG: onset to groin, IA: intra arterial, APT: antiplatelet therapy, eCAS: emergent carotid stenting

## DISCUSSION

In this single-center cohort, eCAS during mechanical thrombectomy was associated with higher rates of successful reperfusion and a more than threefold increase in excellent functional outcomes at 90 days, without an observed increase in sICH or mortality, suggesting a potential efficacy and safety signal of eCAS in carefully selected patients.

Our results are consistent with the growing body of observational evidence suggesting that acute cervical internal carotid artery stenting may improve cerebral reperfusion and facilitate better functional recovery. The TITAN registry—the largest multicenter collaborative analysis of individual patient data across 18 centers—demonstrated that treatment strategies incorporating acute extracranial ICA stenting combined with antithrombotic therapy and intracranial thrombectomy achieved higher recanalization rates and better clinical outcomes without increasing sICH or mortality (5). Similarly, Sivan-Hoffmann et al., in a systematic review and meta-analysis, found that mechanical thrombectomy with concurrent carotid stenting is both feasible and generally safe, with acceptable sICH rates (7%) (7). Data from the STRATIS registry further corroborate these findings, reporting better functional independence among patients receiving acute stenting, with no differences in mortality and sICH but with similar recapitalization rates (6). Comparable observations were reported by Mudassir et al., who found that acute stenting was associated with significantly higher odds of favorable outcome and successful recanalization without increased risk of sICH or mortality (21)

Conversely, some studies have questioned the routine use of eCAS. A meta-analysis by Pires Coelho et al. reported no clinical benefit of acute stenting, noting increased time to recanalization and no significant improvements in successful reperfusion, good functional outcome, or 90-day mortality (1). Similarly, Mitchell et al. did not observe a functional benefit associated with stenting (12). In contrast to our findings, Stampfl et al. reported higher sICH rates in the eCAS group, attributing this to more frequent use =of tirofiban (22). Heck and Brown likewise documented elevated hemorrhagic complications following acute stenting, in their case linked to abciximab administration in an older patient population (23). These discrepancies highlight the heterogeneity of patient selection, procedural techniques, and antithrombotic regimens across studies.

An important and persistent controversy in TLS management is the optimal periprocedural and postprocedural antithrombotic strategy following carotid stenting. In our cohort, patients undergoing eCAS more frequently received tirofiban, dual antiplatelet loading, and dual antiplatelet therapy at discharge. Although these variables introduce possible confounding, the comparable sICH and mortality rates observed between groups suggest that more intensive antiplatelet regimens did not compromise safety and increase effectiveness, similarly to the findings of TITAN registry (5). These findings align with recent observational data suggesting that short-acting glycoprotein IIb/IIIa inhibitors may effectively prevent acute stent thrombosis without substantially increasing the risk of hemorrhage (4). Mudassir et al. also reported no significant differences in sICH between stented and non-stented patients, further supporting the relative safety of tailored antithrombotic strategies(21). Nevertheless, the absence of randomized controlled trials continues to limit definitive conclusions regarding optimal antithrombotic care. The underlying etiology of the cervical lesion is another relevant consideration. In our study, most cervical lesions were due to atherosclerosis (91%), with a minority attributable to dissection (9%). Prior literature suggests that outcomes following stenting are generally similar regardless of etiologic subtype, and our findings are consistent with these observations (24).

This study has several limitations. First, its retrospective, single-center design limits generalizability and carries inherent risks of selection bias. The choice to perform eCAS versus MTa was not randomized and was influenced by operator preference, anatomical considerations, and clinical factors, which may confound comparisons. Second, the eCAS group exhibited a lower baseline NIHSS, as well as a higher proportion of intra-arterial thrombolysis, tirofiban use, and DAPT at discharge. Although analyses were adjusted for age, sex, and baseline NIHSS, the presence of residual confounding cannot be excluded. Third, despite being one of the largest TLS series from Latin America, the overall sample size (n = 111) and the predominance of an atherosclerotic etiology limit statistical power and the generalizability of the findings to other populations or to patients with dissection-related disease. This limitation is particularly relevant for the detection of differences in infrequent outcomes, such as symptomatic intracranial hemorrhage (sICH) or mortality.

## CONCLUSION

In this 10-year single-center experience, emergent carotid artery stenting during mechanical thrombectomy for tandem-lesion anterior circulation strokes was associated with higher rates of successful recanalization and excellent functional outcomes, without an increase in sICH or mortality. These findings support the feasibility and safety of eCAS in appropriately selected patients and contribute valuable evidence to the ongoing debate regarding optimal TLS management. Prospective multicenter studies and randomized clinical trials are warranted to establish standardized guidelines for this complex stroke subtype.

## Ethical approval and informed consent statements

Ethics approval for this study was waived by the Bioethics Committee of Clínica La Sagrada Familia-ENERI on 1st July 2025

## Data availability statement

The datasets generated during and/or analyzed during the current study are available from the corresponding author on reasonable request.

---

**Figure 1.**
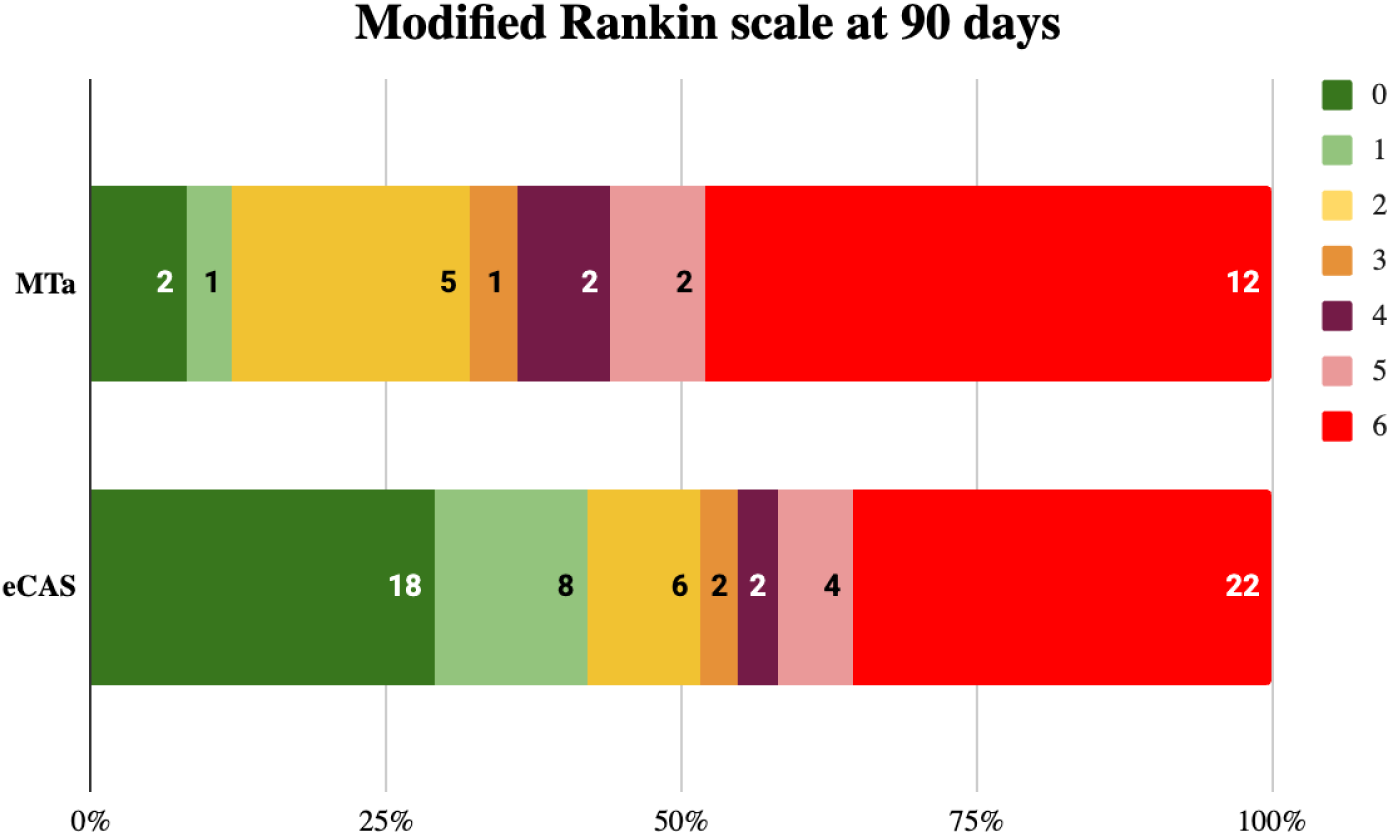
Distribution of 90-day modified Rankin Scale (mRS) Score among patients in eCAS vs MTa groups. *OR* = 2.2; 95% CI 0.93–5.26; *p* .075

